## Supplementary Information for "Sparse haplotype-based fine-scale local ancestry inference at scale reveals recent selection on immune responses"

### 1 Supplementary Note 1

#### 1.1 Genes with shared low LDAS and AAS signals in painting with both 26 populations and 5 continents

Genes with shared low LDAS and AAS signals in painting with both 26 populations and 5 continents can be classified below based on previous findings:
Immune system/response-related genes: **PNRC2** (mRNA degradation), **SRSF10** (RNA splicing), **MRC1** (key role in the binding of glycoproteins)
Development: **PNRC2** (mRNA degradation), **SRSF10** (RNA splicing)
Metabolic diseases: **SRSF10** (RNA splicing), **MRC1** key role in binding of glycoproteins)
Neurological diseases: **SRSF10** (RNA splicing), **DLGAP1** (key role in neurotransmission)
No known disease associations: **TMEM236** (perhaps cancer biomarker)

**PNRC2**

PNRC2 (proline-rich nuclear receptor coregulatory protein 2) is expressed in many immune cells and is involved in nuclear-transcribed mRNA catabolic process, nonsense-mediated decay. It is located in the Golgi apparatus, P-body, and nucleoplasm.

PNRC2 is essential in mammalian nonsense-mediated mRNA decay (NMD), mediating the interaction between the NMD machinery and the decapping complex[1]. It may target the NMD machinery to the P-body and recruit the decapping machinery to aberrant mRNAs, and it is required for UPF1/RENT1 localization to the P-body[1]. It also plays a role in glucocorticoid receptor-mediated mRNA degradation (GR-mediated mRNA decay (GMD)) by interacting with the glucocorticoid receptor NR3C1 in a ligand-dependent manner when it is bound to the 5' UTR of target mRNAs and recruiting the RNA helicase UPF1 and the mRNA-decapping enzyme DCP1A, leading to RNA decay[2]. In addition, it acts as a nuclear receptor coactivator[3]. It may play a role in controlling the energy balance between energy storage and energy expenditure.

GMD functions in the chemotaxis of human monocytes by targeting chemokine (C-C motif) ligand 2 (CCL2) mRNA. The study provides molecular evidence of a posttranscriptional role of the well-studied nuclear hormone receptor, GR, which is traditionally considered a transcription factor[2]. CCL2 is primarily secreted by monocytes, macrophages and dendritic cells. The platelet-derived growth factor is a major inducer of the CCL2 gene.

In addition, CCL2 expression is a prognostic biomarker in hepatocellular carcinoma, breast cancer and bladder cancer.

Diseases associated with PNRC2 include Murray Valley encephalitis ( a severe but rare infection caused by the Murray Valley encephalitis virus. It is spread to humans by infected mosquitoes) and holoprosencephaly 3 (the brain fails to divide correctly into the right and left hemispheres).

**SRSF10**

SRSF10 (Serine And Arginine Rich Splicing Factor 10). The SRSF10 gene product is a member of the serine-arginine (SR) family of proteins involved in constitutive and regulated RNA splicing. The SRSF10 protein is composed of an RNA binding domain (RRM) and arginine and serine-rich auxiliary domains (RS) that guide interactions with other proteins. The phosphorylation status of SRSF10 is of paramount importance for its activity and is subjected to changes during mitosis, heat shock, and DNA damage. SRSF10 overexpression has functional consequences in a growing list of cancers.

By controlling the alternative splicing of specific transcripts, SRSF10 has also been implicated in glucose, fat, and cholesterol metabolism, embryonic heart development, and neurological processes.

SRSF10 is also essential for the proper expression and processing of HIV-1 and other viral transcripts. SRSF10 promotes tumorigenesis of cervix, was upregulated by HPV E6E7 via E2F1 transcriptional activation. SRSF10 modulates the alternate terminator of interleukin-1 receptor accessory protein exon 13 to increase production of the membrane form of interleukin-1 receptor accessory protein. SRSF10-mediated mIL1RAP upregulates the expression of the 'don't eat me' signal CD47 to inhibit macrophage phagocytosis by promoting nuclear factor-$\kappa$B activation, which is pivotal in inflammatory, immune, and tumorigenesis processes[4].

Diseases associated with SRSF10 include alexithymia, alcoholic ketoacidosis, glucose, fat, and cholesterol metabolism aberrations, developmental diseases, neurological diseases, infection (HIV-1 and other viruses), and cancer (e.g., HPV-related cervix cancer).

**TMEM236**

TMEM236 (Transmembrane Protein 236) encodes a protein predicted to be an integral component of membranes. TMEM236 is a novel, not well-examined gene; it is primarily expressed in the intestine and at low levels in the brain. TMEM236 is not expressed in immune system cells[5].

There are no known disease associations yet, but expression is downregulated in colorectal cancers[6]. Moreover, TMEM236 has been found as part of a hybrid RNA transcript (TMEM236-MRC1) in chronic myeloid leukemia (CML) patients and has been suggested to have oncogenic potential[7]. Although TMEM236 has been detected in colorectal cancer, pancreatic cancer, stomach cancer, and leukemia (CML), detection of the expressed gene is, so far, not prognosic in cancers[5].

Expression measurements have been used in a predictive model of immunotherapy efficacy for patients with lung squamous cell carcinoma based on the degree of tumour-infiltrating immune cells in the tumour microenvironment[8].

**MRC1**

MRC1 (Mannose Receptor C-Type 1) is a Protein Coding gene. It is expressed on the surface of macrophages, immature dendritic cells, liver sinusoidal endothelial cells, and the surface of skin cells. It plays a crucial role in the immune system (phagocytosis, antigen presentation, and cytokine release) and in the removal of various waste products (including sulphated glycoprotein hormones).

The mannose receptor binds high-mannose structures on the surface of potentially pathogenic viruses, bacteria, and fungi to neutralise them by phagocytic engulfment. The mannose receptor on liver sinusoidal endothelial cells removes many waste materials ranging from soluble macromolecules to large particulate matter. It plays a role in antigen uptake and presentation by immature dendritic cells in the adaptive immune system. Upon binding to the receptor, mannosylated antigens are internalised and transported to endocytic compartments within the cell for loading onto HLA molecules.

The cytoplasmic tail of the mannose receptor does not contain any signalling motifs. Nevertheless, the receptor has proven to be essential for the production of both pro- and anti-inflammatory cytokines, indicating a more passive role for the receptor in the phagocytosis of pathogens. The mannose receptor is expressed at low levels during inflammation and high levels during the resolution of inflammation to ensure inflammatory agents are removed from the circulation only at the appropriate time. Lastly, the N-terminal cysteine-rich domain of the mannose receptor plays an essential role in the recognition of sulphated glycoprotein hormones and their clearance from circulation.

Diseases associated with MRC1 include infectious diseases (e.g., Dengue virus infection (mediates infection as facilitates virus uptake in macrophages) and (in contrast) HIV-1 (restrictive factor)) and Gaucher's disease (a rare, inherited metabolic disorder in which deficiency of the enzyme glucocerebrosidase results in the accumulation of harmful quantities of lipids, specifically the glycolipid glucocerebroside throughout the body).

#### DLGAP1

DLGAP1 (DLG Associated Protein 1) is a Protein Coding gene.

The postsynaptic density resets the synapse in the nervous system after each synaptic firing. It comprises numerous proteins, including a family of Discs large associated proteins 1, 2, 3 and 4 (DLGAP1-4) that act as scaffold proteins in the postsynaptic density. They link the glutamate receptors in the postsynaptic membrane to other glutamate receptors, to signalling proteins and to cytoskeleton components. With the central localisation in the postsynapse, the DLGAP family plays a vital role in synaptic scaling by regulating the turnover of both ionotropic and metabotropic glutamate receptors in response to synaptic activity. The DLGAP family has been directly linked to a variety of psychological and neurological disorders. A review of DLGAP family is available[9].

Diseases associated with DLGAP1 include retinitis pigmentosa 55 and obsessive-compulsive disorder.

#### 1.2 Genes with shared low LDAS and AAS signals only in painting with 26 populations

Genes with shared low LDAS and AAS signals only in painting with 26 populations can be classified below based on previous findings:
Immune system/response related genes: **HLA-DRB1**, **SIRPD** (expressed in macrophages, neutrophiles and/or T cells in some tissues), **SIRPB1** (autoimmune diseases, infection, innate immune responses, phagocytosis-neural debris)
LncRNA: **NHEG1**, **LINC03004**, **LINC01432**, **SLC39A12-AS1** (cancer, androgenic baldness)
Zink/nucleoside/various transport: **SLC39A12**, **SLC29A3**, **SLC39A12-AS1**, **TMEM241** (neurological diseases, pulmonary hypertension, acrodermatitis, H syndrome, PHID, predicted transport of various)
cell-cell adhesion glycoproteins/ endothelial cell to extracellular matrix adhesion: **CDH23** (deafness), **THSD1** (aneurysm, intracranial bleeding)
Pseudogene: **HLA-DRB1**, **HLA-DRB6**, **PTPN11P3**, **CYP2T3P** (No known disease associations, perhaps T2DM in Sindhi population)
Possible role in reproduction: **SIRPD** (expression enriched in spermatides)
No known disease associations: **PTPN11P3**, **SIRPD**

**HLA-DRB1 and HLA-DRB6**

HLA-DRB6 (Major Histocompatibility Complex, Class II, DR Beta 6) is a Pseudogene. There are no known robust disease associations.

HLA-DRB1 (Major Histocompatibility Complex, Class II, DR Beta 1), is responsible for presenting antigens to T cells and regulating immune responses.

HLA-DRB1 belongs to a group of genes called HLA class II beta chain paralogs. HLA class II molecules are made up of two chains - an alpha chain called DRA and a beta chain called DRB - that are attached to the membrane. The beta chain, which is about 26-28 kDa, is encoded by 6 exons. The first exon encodes the leader peptide, the second and third exons encode the two extracellular domains, the fourth exon encodes the transmembrane domain, and the fifth exon encodes the cytoplasmic tail.

The beta chain contains all the variations that determine which peptides can bind to the molecule. Hundreds of different variations of DRB1 have been identified, some of which are associated with certain diseases or conditions (autoimmune diseases and infection (susceptibility/protection)).

Specific HLA-DRB1 alleles, particularly those encoding the 'shared epitope', such as HLA-DRB104:01, HLA-DRB104:04, HLA-DRB101:01, and others, are associated with an increased risk of developing rheumatoid arthritis. Various HLA-DRB1 alleles, including HLA-DRB103 and HLA-DRB115, have been linked to susceptibility to systemic lupus erythematosus. Variants of HLA-DRB1 have been implicated in autoimmune thyroid diseases like Hashimoto's thyroiditis and

Graves' disease. Certain HLA-DRB1 alleles, such as HLA-DRB103:01 and HLA-DRB103:02, are associated with an increased risk of celiac disease. Some HLA-DRB1 alleles, including HLA-DRB115:01, have been linked to susceptibility to multiple sclerosis.

Specific HLA-DRB1 alleles have shown associations with resistance or susceptibility to HIV infection and progression to AIDS. For example, HLA-DRB113:02 has been associated with slower disease progression, while some alleles like HLA-DRB115:02 have been linked to increased susceptibility. Hepatitis B and C: Specific HLA-DRB1 alleles have been linked to either susceptibility or protection against hepatitis B and C virus infections.

#### NHEG1

NHEG1 (Neuroblastoma Highly Expressed DDX5 Stabilizing LncRNA 1) is an RNA Gene and is affiliated with the lncRNA class. Diseases associated with NHEG1 include Neuroblastoma (NB).

NHEG1 expression is essential for promoting the tumorigenesis and aggressiveness of NB cells. Treatment with siRNAs against NHEG1 or DDX5 reduces the tumour growth and prolongs the survival time of nude mice bearing xenografts.

Long Noncoding RNA NHEG1 Drives b-Catenin Transactivation and Neuroblastoma Progression through Interacting with DDX5.

#### LINC03004 and LINC01432

There are no strong disease associations with LINC03004. It is highly expressed in the testis and gall bladder in healthy individuals. It affects the copy number variation of the genes EYS (retina), NKAIN2 (normal brain), and TARID (TARID (TCF21 Antisense RNA Inducing Promoter Demethylation). Diseases associated with TARID include deafness (autosomal dominant) and dilated cardiomyopathy).

There is a weak association in the Expression Atlas only to non-small cell lung cancer, hepatocellular carcinoma, and breast carcinoma.

Diseases associated with LINC01432 variants include retroperitoneum carcinoma and early-onset androgenetic alopecia.

#### SLC39A12

SLC39A12 (Solute Carrier Family 39 Member 12) belongs to a subfamily of proteins that show structural characteristics of zinc transporters[10]. The SLC39A12 gene encodes the zinc transporter protein ZIP12; it is expressed across many tissues and is highly abundant in the vertebrate nervous system.

Zinc is an essential cofactor for hundreds of enzymes. It is involved in protein, nucleic acid, carbohydrate, and lipid metabolism and in the control of gene transcription, growth, development, and differentiation.

Diseases include acrodermatitis enteropathica, zink-deficiency type, and a study demonstrated that SLC39A12 expression was higher in the dorsolateral prefrontal cortex in individuals with schizophrenia in comparison with controls[11]. Given that ZIP12 is highly expressed in the brain and that susceptibility-weighted MRI is associated with brain metal content, ZIP12 may affect neurological diseases and psychiatric illnesses such as Parkinson's disease, Alzheimer's disease, and schizophrenia.

Outside the nervous system, hypoxia induces ZIP12 expression in multiple mammalian species, including humans, which leads to endothelial and smooth muscle thickening in the lung and contributes to pulmonary hypertension. Studies have also associated ZIP12 with diseases such as cancer. A comprehensive review for SLC39A12 is available[12].

#### SLC29A3

SLC29A3 (Solute Carrier Family 29 Member 3) encodes a nucleoside transporter called equilibrative nucleoside transporter 3 (ENT3). ENT3 is found in the membranes surrounding cell structures known as lysosomes and mitochondria and transports nucleosides generated by the breakdown of DNA and RNA out of lysosomes into the cell so they can be reused. Thus, ENT3 plays a role in cellular uptake of nucleosides, nucleobases, and their related analogs.

Mutations in this gene have been associated with histiocytosis-lymphadenopathy plus (H) syndrome, which is characterized by cutaneous hyperpigmentation and hypertrichosis, hepatosplenomegaly, heart anomalies, and hypogonadism.

A related disorder, pigmented hypertrichosis with insulin-dependent diabetes mellitus (PHID), has also been associated with mutations at this locus. Alternatively spliced transcript variants have been described.

How diminished/lack of ENT3 function causes these diseases is currently unknown.

#### SLC39A12-AS1

SLC39A12-AS1 (SLC39A12 Antisense RNA 1) is an RNA Gene and is affiliated with the lncRNA class.

Function not described but perhaps it has a role in regulating SLC39A12.

#### CDH23

CDH23 (Cadherin Related 23) is a member of the cadherin superfamily, whose genes encode calcium dependent cell-cell adhesion glycoproteins. The encoded protein is thought to be involved in stereocilia organization and hair bundle formation.

The gene is located in a region containing the human deafness loci DFNB12 and USH1D.

Usher syndrome 1D (a congenital, bilateral, profound sensorineural hearing loss, vestibular areflexia, with adolescent-onset retinitis pigmentosa (RP)(vision impairment/loss)) and nonsyndromic autosomal recessive deafness DFNB12 are caused by allelic mutations of this cadherin-like gene. Upregulation of this gene may also be associated with breast cancer.

#### TMEM241

TMEM241 (Transmembrane Protein 241) is likely to be expressed in all tissues at varying levels from basal to moderate expression.

It is predicted to be involved in antiporter activity as well as carbohydrate and transmembrane transport and is also predicted to be an integral component of the Golgi apparatus membrane.

Some studies have found changes in the expression of TMEM241, e.g., in cases of acute megakaryoblastic leukaemia, TMEM241 was found to be one of the most upregulated genes, but the pathogenic importance is uncertain.

#### PTPN11P3

PTPN11P3 (Protein Tyrosine Phosphatase Non-Receptor Type 11 Pseudogene) which is close to LINC03004 and also has very high African ancestry.

There are no known disease associations.

#### CYP2T3P

CYP2T3P (Cytochrome P450 Family 2 Subfamily T Member 3, Pseudogene) is a Pseudogene.

The only disease association is with type 2 diabetes mellitus in 28 pooled whole blood genomes of 1402 participants from the Diabetes In Sindhi Families In Nagpur (India) (DISFIN) study, but I am not sure the study is robust[13].

#### SIRPD

SIRPD (Signal Regulatory Protein Delta) is predicted to be located in the extracellular region and expected to be active in the plasma membrane. An important paralog of this gene is SIRPG. SIRPD is expressed in immune cells (macrophages, neutrophiles, and/or T cells) in certain tissues and testis (early and late spermatides[14]).

There are no known disease associations.

#### SIRPB1

SIRPB1 (Signal Regulatory Protein Beta 1) is a signal-regulatory-protein (SIRP) family member and belongs to the immunoglobulin superfamily. SIRP family members are receptor-type transmembrane glycoproteins known to be involved in the negative regulation of receptor tyrosine kinase-coupled signalling processes. This protein interacted with TYROBP/DAP12, a protein-bearing immunoreceptor tyrosine-based activation motifs. This protein was also reported to participate in the recruitment of tyrosine kinase SYK.

SIRPB1 is a novel IFN-induced microglial receptor that supports the clearance of neural debris and amyloid-beta aggregates by stimulating phagocytosis.

Multiple transcript variants encoding different isoforms have been found for this gene.

SIRPB1 is involved in immune responses (likely primarily innate immune responses), including stimulating phagocytosis.

Disease associations include polycystic lipomembranous osteodysplasia with sclerosing leukoencephalopathy 1. Moreover, a rare gain-of-function frameshift variant in SIRPB1 is associated with Han Chinese patients with Crohn's disease (CD)[15].

**THSD1**

THSD1 (Thrombospondin Type 1 Domain Containing 1) encodes a protein that contains a type 1 thrombospondin domain, which is found in several proteins involved in the complement pathway, and in extracellular matrix proteins. Among its related pathways are O-linked glycosylation of mucins and metabolism of proteins. Alternatively, spliced transcript variants encoding different isoforms have been observed for this gene.

Disease associations include aneurysm, intracranial berry 12, and lymphatic malformation 13.

A study[16] found THSD1 mutations in familial and sporadic IA (ruptured intracranial aneurysm) patients and shows that THSD1 loss resulted in cerebral bleeding in 2 animal models. This finding provides insight into IA and subarachnoid hemorrhage pathogenesis and provides a new understanding of THSD1 function, including endothelial cell to extracellular matrix adhesion.

#### 1.3 Genes with shared low LDAS but without AAS signals in painting with either 26 populations or 5 continents

Genes with shared low LDAS but without AAS signals in painting with either 26 populations or 5 continents can be classified below based on previous findings:

Immune system/response-related genes: **TRBV4-1**, **TRBV6-8**, **TRBV6-6**, **TRBV12-2**, **TRBV11-1**, **TRBV10-1**, **TRBV23-1**, **TRBV24-1**, **IL20RA**, **SIRPG**, **SIRPG-AS1**, **TMPRSS11E** (infections, autoimmune diseases)

Regulation of the cell cycle - RNA metabolism – development: **CABLES1**, **YTHDC1** (cancers)

Detoxification: **UGT2B15** (autoimmune diseases, cancers)

**TRBV genes**

TRBV4-1: Some studies have suggested a potential association between TRBV4-1 and protection against infections caused by specific pathogens, such as certain viral infections (e.g., human immunodeficiency virus (HIV)[17], influenza viruses[18]), herpesviruses (e.g. CMV[19]), hepatitis C virus (HCV)[20]) and cancer[21,22,23] (only in 26-pop painting).

TRBV6-8: Research has explored associations between TRBV6-8 and protection against viral infections, including HCV[24], SARS-CoV-2[25], some herpes viruses and cancer[21]. The TCR$\beta$ repertoire of MAIT cells appears to be biased towards TRBV-6 families. MAIT cells are a type of immune cell that respond to various microorganisms such as Gram-positive bacteria, Gram-negative bacteria, and yeasts. These cells have the ability to fight against infections caused by these microorganisms[26].

TRBV6-6: Associations have been investigated between TRBV6-6 and protection against certain viral infections (e.g.influenza[27] and HIV)[21,28,17], and tuberculosis[29]. The TCR$\beta$ repertoire of MAIT cells appears to be biased towards TRBV-6 families. MAIT cells are a type of immune cell that respond to various microorganisms such as Gram-positive bacteria, Gram-negative bacteria, and yeasts. These cells have the ability to fight against infections caused by these microorganisms[26].

TRBV12-2: Some research has suggested associations between TRBV12-2 and increased risk for cerebral malaria despite TRBV12-2 being a pseudogene[30].

TRBV11-1: This gene segment has been studied in the context of protection against viral infections like HIV (Human Immunodeficiency Virus), acute hepatitis B infection[31], and certain herpesviruses. The TRBV 5, 10, and 11 gene families are preferentially expanded in HIV-1 infection when the HSK epitope is presented by HLA-B*08:01.

TRBV10-1: Associations have been investigated between TRBV10-1 and protection against infections caused by certain viruses (possibly including SARS-CoV-2[32]) and intracellular bacteria and cancer. The TRBV 5, 10, and 11 gene families are preferentially expanded in HIV-1 infection when the HSK epitope is presented by HLA-B*08:01.

TRBV23-1: Studies have explored associations between TRBV23-1 and protection against viral infections. Specific viral pathogens involved are still under investigation. TRBV23-1 is expressed on Natural Killer T (NKT) cells[33].

TRBV24-1: Research has looked into associations between TRBV24-1 and protection against certain viral infections including herpes simplex type 1 and acute hepatitis B infection[31,34], and cancer[21].

In humans, specific TRBV (T-cell receptor beta variable) gene expression can vary widely among individuals and across different T-cell populations. However, specific TRBV genes tend to be more frequently expressed or prevalent in the T-cell receptor repertoire across the general population.

While the exact ranking or prevalence of TRBV gene expression can fluctuate based on factors like age, health status, and antigen exposure, some TRBV genes have been observed to be relatively more commonly expressed than others. None of the most common TRBV are included.

The list is noteworthy for its multiple associations with specific pathogens (especially ancient (herpesviruses, bacteria) and old (HBV, HCV, influenza)), bacteria, and non-specific pathogens.

## IL20RA

IL20RA, also known as Interleukin-20 Receptor Alpha, is a part of the interleukin-20 receptor complex, which includes IL20RB. IL20RA plays a significant role in the signalling pathway of IL-20 cytokines as it mediates the pro-inflammatory effects of IL-20 cytokines. These cytokines are a part of the IL-10 cytokine family and have been linked to various inflammatory and autoimmune diseases (including psoriasis, rheumatoid arthritis, inflammatory bowel disease, atopic dermatitis, and ankylosing spondylitis). The signalling pathway through IL20RA helps to regulate immune responses, tissue homeostasis, and inflammation. It is a central player in the immune system.

#### SIRPG and SIRPG-AS1

SIRPG is a member of the signal-regulatory protein (SIRP) family and also belongs to the immunoglobulin superfamily. SIRP family members are receptor-type transmembrane glycoproteins known to be involved in the negative regulation of receptor tyrosine kinase-coupled signalling processes. Alternatively, spliced transcript variants encoding different isoforms have been described.

Multiple GWAS studies have shown that the SNP rs2281808 TT variant within the SIRPG gene is associated with autoimmune diseases, such as type 1 diabetes.

A recent study investigated SIRPG genotypes and their effects on the fate and function of human T-cells[35]. They found that the presence of the T variant resulted in a reduction of the signal regulatory protein gamma (SIRP$\gamma$) expression on T-cells. SIRP$\gamma$ is a transmembrane protein found on immune cells that helps to regulate immune responses, cell adhesion, and phagocytosis.

SIRPD (Signal Regulatory Protein Delta) is a protein-coding gene. An important paralog of this gene is SIRPG. It is predicted to be located in the extracellular region and to be active in the plasma membrane.

SIRPG-AS1 (only 26-pop painting) is a long non-coding RNA (lncRNA) that is produced from the opposite strand of the SIRPG gene locus. It likely plays a key role in regulating gene expression and cellular processes. SIRPG-AS1 may control the expression of SIRPG, the gene that encodes the signal regulatory protein gamma (SIRP$\gamma$). SIRP$\gamma$ is a transmembrane protein found on immune cells that helps to regulate immune responses, cell adhesion, and phagocytosis.

#### TMPRSS11E

TMPRSS11E is a gene that belongs to the type II transmembrane serine protease family. This protease plays a crucial role in the activation of other proteins, including protease-activated receptors (PARs) and viral glycoproteins. TMPRSS11E is involved in various important physiological processes, such as epithelial barrier function, inflammation and wound healing.

Dysregulation of TMPRSS11E expression or activity has been associated with inflammatory disorders such as asthma, chronic obstructive pulmonary disease (COPD), and inflammatory bowel disease (IBD).

#### CABLES1

CABLES1 (Cdk5 And Abl Enzyme Substrate 1) encodes a protein involved in the regulation of the cell cycle through interactions with several cyclin-dependent kinases.

One study[36] reported aberrant splicing of transcripts from this gene, which results in the removal of the cyclin binding domain only in human cancer cells, and another study found a reduction in gene expression in colorectal cancers[37], while links to pituitary cancer have been observed. Other disease associations are e.g., endometrial hyperplasia.

Multiple transcript variants encoding different isoforms have been found for this gene.

#### YTHDC1

YTHDC1 (YTH Domain Containing 1) is a gene that encodes a protein belonging to the YTH domain-containing protein family. The YTH domain has a specific binding ability to N6-methyladenosine (m6A), which is a common modification found in mRNA. YTHDC1 is involved in various RNA metabolism activities, such as mRNA splicing, mRNA export, and RNA stability, and has been linked to germ cell development, meiosis, and embryogenesis.

Mutations in the YTHDC1 gene have been associated with infertility and developmental disorders. YTHDC1 is associated with triple negative breast cancer (TNBC) (including key player in metastasis), endometrial cancer, glioblastoma, and acute myeloid leukemia (AML) progression.

#### UGT2B15

The UGT2B15 gene is responsible for encoding a member of the UDP-glucuronosyltransferase (UGT) enzyme family. These enzymes play a crucial role in the detoxification and elimination of various substances, including drugs, xenobiotics, and endogenous compounds, through a process known as glucuronidation. UGT2B15 is primarily expressed in the liver and is responsible for glucuronidating different substrates like bile acids, steroids, and environmental toxins. Additionally, genetic variations in the UGT2B15 gene can affect drug metabolism and make individuals more susceptible to certain diseases.

Diseases associated with UGT2B15 include Anxiety and Crigler-Najjar Syndrome, Type I. It is also involved in drug metabolism in the liver and may be linked to liver diseases like drug-induced liver injury or liver cirrhosis. The UGT2B15 gene is negatively regulated in castration-resistant prostate cancer and lymph node metastases.

UGT2B15-mediated metabolism is believed to play a role in inflammatory diseases like inflammatory bowel disease (IBD) and asthma. Therefore, variations in the UGT2B15 gene may affect susceptibility to these conditions or response to treatment.

#### 1.4 Genes with shared AAS signals but without LDAS signals in painting with either 26 populations or 5 continents

Genes with shared AAS signals but without LDAS signals in painting with either 26 populations or 5 continents can be classified below based on previous findings:
Immune response: **CNR2** (likely inflammation), **STAM**, **PDPK1**, **RPL34P29**
Infection: **STAM** (facilitates dengue virus entry), **PDPK1**
Collagen (skin/connective tissue/mucous membrane) diseases: **NOMO2**
Nervous system: **FAM30B** (Charcot–Marie–Tooth disease), **CNR2** (effects of marijuana), **ASAN**, **STAM** (Charcot–Marie–Tooth disease, and demyelination), **PDPK1**, **RPL34P29** (possible link to age of Alzheimer disease onset)
Psychiatric diseases: **CNR2** (eating disorder)
Cardiovascular system: **FAM30B** (Fanconi anaemia)
Development: **NOMO2**, **ASNS**, **MKKS** (essential to cytokinesis), **SLX4IP** (telomere maintenance mechanisms), **PDPK1**
Vesicular transport: **ASAP2** (to Golgi), **MKKS** (transport of vesicles to the cilia)
Vitamin-dependent: **ASAP2** (vitamin D)
Metabolism: **CYP2F1**
Cancer: **ASAP2**, **ABCC6P1**, **TPTE2P2** (only possibility), **SLX4IP** (telomere maintenance mechanisms), **PDPK1**
LncRNA: **FAM30B**, **ABCC6P1**, **LINC01674**
Pseudogene: **NCOA5LP**, **ABCC6P1**, **BTBD6P1**, **TPTE2P2**, **CNN2P7**, **RPL34P29**
No known disease associations: **FAM30B**, **NCOA5LP**, **CYP2F1**

The associations for those genes are more diverse, and they include many LncRNA and pseudogenes. There are two genes associated with a rare genetic disorder (Charcot–Marie–Tooth disease), but also have other associations. Facilitation of dengue virus infection by **STAM** (the facilitation of dengue virus infection also exists in MRC1 which has both AAS and low LDAS signals). Together, this might suggest that dengue infection was fairly localised in the past and that selection against variants that facilitate infection did not occur in many populations due to a lack of exposure to the virus, which is in line with epidemiological data[38].

**ASAP2**
ASAP2 (ArfGAP With SH3 Domain, Ankyrin Repeat And PH Domain 2). ASAP2 encodes a multidomain protein containing an N-terminal alpha-helical region with a coiled-coil motif, followed by a pleckstrin homology (PH) domain, an Arf-GAP domain, an ankyrin homology region, a proline-rich region, and a C-terminal Src homology 3 (SH3) domain. The protein localizes in the Golgi apparatus and at the plasma membrane, where it colocalizes with protein tyrosine kinase

2-beta (PYK2). The encoded protein forms a stable complex with PYK2 in vivo. This interaction appears to be mediated by the binding of its SH3 domain to the C-terminal proline-rich domain of PYK2. The encoded protein is tyrosine phosphorylated by activated PYK2. It has catalytic activity for class I and II ArfGAPs in vitro, and can bind the class III Arf ARF6 without immediate GAP activity.

The encoded protein is believed to function as an ARF GAP that controls ARF-mediated vesicle budding when recruited to Golgi membranes. In addition, it functions as a substrate and downstream target for PYK2 and SRC, a pathway that may be involved in the regulation of vesicular transport. Multiple transcript variants encoding different isoforms have been found for this gene.

ASAP2 is a primary 1,25(OH)2D3 target gene in monocytes and macrophages[39]. In human monocytes (THP-1 cells) the ASAP2 gene is more weakly expressed but more and faster inducible by the biologically active form of vitamin D, 1$\alpha$,25-dihydroxyvitamin D3 (1,25(OH)2D3), than in M2-type macrophages (phorbol ester-differentiated THP-1 cells). Within the investigated genomic region, the basal mRNA expressions of the neighbouring genes are comparably high in both monocytes and macrophages, but the ASAP2 gene is the only primary 1,25(OH)2D3 target[39].

Disease associations: cancer, driver of cancer progression. Clinical analysis with PDAC datasets showed that ASAP2 was overexpressed in Pancreatic ductal adenocarcinoma (PDAC) cells based on increased DNA copy numbers, and high ASAP2 expression contributed to a poor prognosis in PDAC[40].

A study highlights the importance of ASAP2 in sustaining c-MET signalling, which can facilitate hepatocellular carcinoma (HCC) progression[41].

#### FAM30B

FAM30B (Family With Sequence Similarity 30 Member B) is an lncRNA. Normal function is as yet undefined.

Disease association is uncertain as FAM30 can be divided into FAM30A, B, or C and papers usually just use FAM30.

FAM30 was implicated in one male with Charcot–Marie–Tooth disease (CMT) and the symptoms of sorbitol dehydrogenase deficiency with peripheral neuropathy (AR). CMT is a genetically and clinically heterogeneous group of disorders characterized by progressive distal limb weakness, gait disturbance and sensorimotor polyneuropathy[42].

FAM30 was implicated in some cases of Fanconi anaemia (FA). FA is a rare recessive disease resulting from mutations in one of at least 16 different genes[43].

#### NOMO2

NOMO2 (NODAL Modulator 2) encodes a protein originally thought to be related to the collagenase gene family. This gene is one of three highly similar genes in a region of duplication located on the p arm of chromosome 16. These three genes encode closely related proteins that may have the same function. The protein encoded by one of these genes has been identified as part of a protein complex that participates in the Nodal signalling pathway (a signal transduction pathway important in regional and cellular differentiation) during vertebrate development.

Mutations in the ABCC6 transporter gene (see ABCC6P1 later), which is located nearby, rather than mutations in this gene are associated with pseudoxanthoma elasticum (PXE) (Elastic tissue in the body becomes mineralized as calcium is deposited in the tissue). Two transcripts encoding different isoforms have been described.

Diseases associated with NOMO2 include (perhaps to some extent) Pseudoxanthoma Elasticum and Noma. Noma is a rapidly progressing severe gangrenous disease of the mouth and the face. It mostly affects children aged 2–6 years suffering from malnutrition, affected by infectious diseases, living in extreme poverty with poor oral health or with weakened immune systems.

Component of the multi-pass translocon (MPT) complex that mediates insertion of multi-pass membrane proteins into the lipid bilayer of membranes[44,45].

#### NCOA5LP

NCOA5LP (Nuclear Receptor Coactivator 5 Like, Pseudogene) is a pseudogene. Normal function is as yet undefined.

There are no known disease associations.

#### ABCC6P1

ABCC6P1 (ATP Binding Cassette Subfamily C Member 6 Pseudogene 1).

Chromosome 16 locates lncRNA ABCC6P1, which, along with its parent ABCC6, shares near-identical promoter sequences and similar tissue-specific expression profiles. In addition, ABCC6P1 is transcriptionally active and can regulate ABCC6 at the transcriptional level.

Disease associations: cancer. A study found that ABCC6P1 was upregulated in thyroid cancer and that it promoted the proliferation and migration of papillary thyroid cancer cells via the Wnt/$\beta$-catenin signalling pathway[46].

#### CNR2

CNR2 (Cannabinoid Receptor 2 (Macrophage)). The proteins encoded by CNR2 and the cannabinoid receptor 1 (brain) (CNR1) gene have the characteristics of a guanine nucleotide-binding protein (G-protein)-coupled receptor for cannabinoids (the principal psychoactive ingredient of marijuana). They inhibit adenylate cyclase activity in a dose-dependent, stereoselective, and pertussis toxin-sensitive manner.

These proteins are involved in the cannabinoid-induced CNS effects (including alterations in mood and cognition) experienced by users of marijuana. CNR2 may function in inflammatory response, nociceptive transmission, and bone homeostasis.

Disease associations: Purulent labyrinthitis, eating disorders, and plays a central role in renal tubular mitochondrial dysfunction and kidney ageing.

#### BTBD6P1

BTBD6P1 (BTB Domain Containing 6 Pseudogene 1) is a pseudogene.

There are no known function or disease associations yet.

#### ASNS

ASNS (Asparagine Synthetase (Glutamine-Hydrolysing) encodes an enzyme synthesising asparagine. The enzyme is found in cells throughout the body, where it converts the protein building block (amino acid) aspartic acid to the amino acid asparagine. Another amino acid, glutamine, helps in the conversion and is itself converted to the amino acid glutamic acid during the process. It is thought that asparagine synthetase helps to maintain the normal balance of these four amino acids in the body.

Disease associations: Asparagine synthetase deficiency is a condition that causes neurological problems in affected individuals starting soon after birth. Most people with this condition have an unusually small head size (microcephaly) that worsens over time due to loss (atrophy) of brain tissue. It also causes neuronal migration disorders (NMDs). NMDs are a group of congenital disabilities caused by the abnormal migration of neurons (nerve cells) in the developing brain and nervous system.

#### STAM

STAM (Signal Transducing Adaptor Molecule) mediate downstream signalling of cytokines and growth factors. Upon IL-2 and GM-CSL stimulation, it plays a role in signalling, leading to DNA synthesis and MYC induction.

It may also play a role in T-cell development. It is involved in the down-regulation of receptor tyrosine kinase via the multivesicular body (MVBs) when complexed with HGS (ESCRT-0 complex). The ESCRT-0 complex binds ubiquitin and acts as a sorting machinery that recognizes ubiquitinated receptors and transfers them to further sequential lysosomal sorting/trafficking processes.

It may also play a role in ER to Golgi trafficking by interacting with the coat protein II complex. Alternatively, spliced transcript variants have been observed for this gene

Disease associations: Dengue virus infection (facilitates virus entry), Charcot-Marie-Tooth disease, and demyelination, Type 1C.

STAM1 mRNA expression is associated with survival in clear cell renal cell carcinoma (ccRCC).

#### TPTE2P2

TPTE2P2 (TPTE2 Pseudogene 2) is a pseudogene.

Possible driver in aggressive metastasis-prone Lung Cancers: TPTE encodes a testis-specific protein with high sequence similarity to the tumour suppressor PTEN, and TPTE2P2 encodes a testis-specific TPTE pseudogene. It is possible that the ectopic activation of TPTE or a truncated protein produced by TPTE2P2 could interfere with the tumour suppressor activity of PTEN and act as a dominant negative factor with oncogenic activity[47].

There is no known effect and no known robust disease associations.

#### PDPK1

The PDPK1(3-Phosphoinositide Dependent Protein Kinase 1) (also known as PDK1) gene encodes a serine/threonine-protein kinase that plays a central role in regulating cell growth, survival, and proliferation. PDPK1 is a master kinase, phosphorylating and activating, for example, PKB/Akt (a serine/threonine-specific protein kinase that plays a critical role in controlling the balance between survival and death pathways in cells), S6K (which interacts with the translation initiation factor eukaryotic initiation factor 4B), and RSK (a protein kinase involved in signal transduction).

Mice lacking PDPK1 die during early embryonic development, which suggests that PDPK1 is critical for transmitting the growth-promoting signals necessary for normal mammalian development[48].

Disease associations:

Cancer: PDPK1 expression has been found to play a role in several cancers as it mediates cancer cell survival via activation of predominantly the serum/glucocorticoid-regulated kinase 3 (SGK3). Affected cancers include prostate cancer[49], glioma progression (as PDPK1 promotes the proliferation and inhibits the apoptosis of glioma cells)[50], breast[51], and ovarian tumours[52].

Neurological diseases: PDPK1 might play a role in the development of Alzheimer's disease[53].

Infection: PDK1 activity is increased in prion-infected neurons[53].

Immunity: PDK1 is a crucial regulator of immune cell development, connecting PI3K to downstream AKT signalling[54].

#### RPL34P29

RPL34P29 (Ribosomal Protein L34 Pseudogene 29) is a processed pseudogene.

Disease associations:

Immunity: Mutations in RPL34P29 have been implicated in cold medicine-related Stevens-Johnson syndrome/toxic epidermal necrolysis (SJS/TEN) with severe ocular complications[55,56] and the number of common colds[57].

Neurological diseases: Mutations in RPL34P29 have been linked to the age of onset of Alzheimer's disease[58].

#### CYP2F1

CYP2F1 (cytochrome P450 family 2 subfamily F member 1). CYP2F1 encodes a member of the cytochrome P450 superfamily of enzymes. The cytochrome P450 proteins are monooxygenases that catalyse many reactions in drug metabolism and synthesise cholesterol, steroids and other lipids. This protein localizes to the endoplasmic reticulum and is known to dehydrogenate 3-methylindole, an endogenous toxin derived from the fermentation of tryptophan, as well as xenobiotic substrates such as naphthalene and ethoxycoumarin. This gene is part of a large cluster of cytochrome P450 genes from the CYP2A, CYP2B and CYP2F subfamilies on chromosome 19q.

The CYP2F1 is a human cytochrome P450 that is selectively expressed in lung tissue and involved in the metabolism of various pneumotoxicants with potential carcinogenic effects[59].

There are no robust disease associations. A small study demonstrated a link to chronic obstructive pulmonary disease in Taters[60]. CYP2F1 is a quite polymorphic gene, but no specific polymorphisms have been associated with lung cancer (Europe)[61] or nasopharyngeal carcinoma (China)[62].

#### MKKS

MKKS (MKKS Centrosomal Shuttling Protein). MKKS encodes a protein that shares a sequence similarity with other type II chaperonin family members. The encoded protein is a centrosome-shuttling protein and is essential to cytokinesis. This protein also interacts with other type II chaperonin members to form a complex known as the BBSome, which involves ciliary membrane biogenesis and regulates the transport of vesicles to the cilia[63]. This protein is encoded by a downstream open reading frame (dORF). Several upstream open reading frames (uORFs) have been identified, which repress the translation of the dORF, and two of which can encode small mitochondrial membrane proteins. MKKS is probably also a molecular chaperone that assists in the folding of proteins upon ATP hydrolysis[63]. It may play a role in protein processing in limb, cardiac and reproductive system development. Alternative splicing results in multiple transcript variants.

Disease associations: Mutations in MKKS have been observed in patients with Bardet-Biedl syndrome type 6 and McKusick-Kaufman syndrome.

Clinical characteristics: Mutations in the MKKS gene cause Bardet–Biedl syndrome (BBS), a genetically heterogeneous disorder with pleiotropic symptoms. BBS is a rare syndrome characterized by early-onset obesity, rod–cone dystrophy, dyslexia, learning disabilities, postaxial polydactyly, hypogonadism and progressive renal disease. The McKusick–Kaufman syndrome is an allelic form of BBS. However, little is known about how MKKS mutations lead to disease[64].

McKusick-Kaufman syndrome (MKS) is characterized by the combination of postaxial polydactyly (PAP), congenital heart disease (CHD), and hydrometrocolpos (HMC) in females and genital malformations in males (most commonly hypospadias, cryptorchidism, and chordee). Inheritance of MKS follows an autosomal recessive pattern[65].

**SLX4IP**

SLX4IP (SLX4 Interacting Protein) is a protein-coding gene that helps govern telomere maintenance mechanisms (TMMs).

Diseases associated with SLX4IP include myasthenic syndrome, congenital, 18 and Huntington disease-like 1 and cancer (Androgen Receptor-Independent Castration-Resistant Prostate Cancer, breast cancer metastasis, Acute lymphocytic leukemia (ALL) predominance in boys).

Cancer associations in detail: SLX4IP Promotes Telomere Maintenance in Androgen Receptor-Independent Castration-Resistant Prostate Cancer through ALT-like Telomeric PML Localization.

SLX4IP and telomere dynamics dictate breast cancer metastasis and therapeutic responsiveness[66].

The repeatedly detected association of SLX4IP deletion with male sex and the extension of the sex bias to deletion of the TAL1 locus suggest that differential illegitimate V(D)J-mediated recombination events at specific loci may contribute to the consistent observation of higher incidence rates of childhood ALL in boys compared with girls[67].

**LINC01674**

LINC01674 (Long Intergenic Non-Protein Coding RNA 1674) is a lncRNA.

LINC01674 is known to be under selection[68]. The positive and negative iHS values were considered to capture ancient and recent signals of selection, respectively. In total, 17 candidate genomic regions under selection that produced clusters of markers with outlier values were identified, including LINC01674.

There are no known disease associations.

**CNN2P7**

CNN2P7 (Calponin 2 Pseudogene 7) is a pseudogene.

There are no known disease associations.

### 2 Supplementary Note 2

#### 2.1 E-M algorithm with normalised forward and backward probabilities

In Methods section, we have shown the computation of the normalised $\mathbf{f}_j$ and $\mathbf{b}_j^T$ as $\check{\mathbf{f}}_j$ and $\check{\mathbf{b}}_j^T$:

$$\check{\mathbf{f}}_j = \frac{1}{F_j}\mathbf{v}_j \circ \left(\rho_{j-1}\check{\mathbf{f}}_{j-1} + \tilde{\rho}_{j-1}\mathbf{1}_N\right) = \frac{1}{F_j}\check{\mathbf{f}}_j^* = \frac{1}{\prod_{k=1}^{j} F_k}\mathbf{f}_j \tag{1}$$

where $F_j = \sum_{i=1}^{K} \check{f}_{ij}^{*}$

$$\check{\mathbf{b}}_j^T = \frac{1}{B_j}\left(\rho_j \check{\mathbf{d}}_{j+1}^T + \tilde{\rho}_j \check{\check{d}}_{j+1} \mathbf{1}_N^T\right) = \frac{1}{B_j}\check{\mathbf{b}}_j^{T*} = \frac{1}{\prod_{k=j}^{K} B_k}\mathbf{b}_j^T \tag{2}$$

where $\check{\mathbf{d}}_j^T = \mathbf{v}_j \circ \check{\mathbf{b}}_j$ and $\check{\check{d}}_j = \sum_{i=1}^{N} v_{ij}\check{b}_{ij}$.

We can estimate $\lambda$, $\mu$ and the expected length of copied chunks $\hat{l}_i$ using Expectation-Maximisation (E-M) algorithm based on the normalised $f$ and $b$.

Let $\hat{u}_{i,j}$ denote the probability that a target haplotype is copying from the $i$th reference sample at the $j$th SNP given at least one 'switch' has occurred between SNP $j-1$ and $j$.

$$\begin{aligned}\hat{u}_{i,j} &= \frac{1}{P_r(D)}\left[f_{i(j+1)}b_{i(j+1)} - f_{ij}b_{i(j+1)}V_{i(j+1)}\rho_j\right] \\ &= \frac{1}{\prod_{k=1}^{K} F_k}\left[\check{f}_{i(j+1)}\check{b}_{i(j+1)}(\prod_{k=1}^{j+1} F_k)(\prod_{k=j+1}^{K} B_k) - \check{f}_{ij}\check{b}_{i(j+1)}(\prod_{j=1}^{j} F_j)(\prod_{k=j+1}^{K} B_k)V_{i(j+1)}\rho_j\right]\end{aligned} \tag{3}$$

so

$$\begin{aligned}\tilde{u}_j &= \sum_{i=1}^{N} \hat{u}_{i,j} \\ &= \frac{1}{\prod_{k=1}^{K} F_k}\left[\check{\mathbf{f}}_{j+1} \circ \check{\mathbf{b}}_{j+1}(\prod_{k=1}^{j+1} F_k)(\prod_{k=j+1}^{K} B_k) - \check{\mathbf{f}}_j \circ \check{\mathbf{b}}_{j+1} \circ \mathbf{v}_{j+1}(\prod_{k=1}^{j} F_k)(\prod_{k=j+1}^{K} B_k)\rho_j\right]\mathbf{1}_N^T \\ &= \left[a_j^l \check{\mathbf{f}}_{j+1} \circ \check{\mathbf{b}}_{j+1} - a_j^r \check{\mathbf{f}}_j \circ \check{\mathbf{b}}_{j+1} \circ \mathbf{v}_{j+1}\rho_j\right]\mathbf{1}_N^T\end{aligned} \tag{4}$$

where

$$a_j^l = \exp\left(\log(\prod_{k=1}^{j+1} F_k) + \log(\prod_{k=j+1}^{K} B_k) - \log(\prod_{k=1}^{K} F_k)\right)$$

and

$$a_j^r = \exp\left(\log(\prod_{k=1}^{j} F_k) + \log(\prod_{k=j+1}^{K} B_k) - \log(\prod_{k=1}^{K} F_k)\right)$$

In calculation, we use logarithm-expectation transformation to compute $a_j^l$ and $a_j^r$ to avoid the cumulative multiplication of $F_k$ or $B_k$ decimal precision limitation in C++.

#### 2.2 Estimation of recombination scaling constant $\lambda$ using E-M algorithm

To estimate $\lambda$ based on E-M algorithm, we start with an initial value of $\lambda = 400000/N$, and at each simulation, we replace $\lambda$ with

$$\begin{aligned}\lambda^* &= \frac{\sum_{j=1}^{K-1}\left(\left[\sum_{i=1}^{N}\hat{u}_{i,j}\right]\lambda g_j/\left[1.0-\rho_j\right]\right)}{\sum_{j=1}^{K-1} g_j} \\ &= \frac{\sum_{j=1}^{K-1}\left(\tilde{u}_j \lambda g_j/\left[1.0-\rho_j\right]\right)}{\sum_{j=1}^{K-1} g_j},\end{aligned} \tag{5}$$

where each $\tilde{u}_j$ and $\rho_j$ are calculated using the previous estimate of $\lambda$.

**Using E-M to estimate mutation rate $\mu$**

Watterson's estimate can help fix the mutation parameter $\mu$[69]:

$$\mu = \frac{1}{2} \frac{\left(\sum_{k=1}^{N} 1/k\right)^{-1}}{N + \left(\sum_{k=1}^{N} 1/k\right)^{-1}} \tag{6}$$

To use E-M algorithm to estimate $\mu$, we start from Watterson's estimate, and at each simulation we replace $\mu$ with

$$\begin{aligned}
\mu^* =& \frac{\sum_{j=1}^{K} \left(\sum_{i=1}^{N} f_{ij} b_{ij} (1 - M_{ij}) / \Pr(D)\right)}{K} \\
=& \frac{\sum_{j=1}^{K} \left(\sum_{i=1}^{N} \left[\check{f}_{ij} \check{b}_{ij} (\prod_{k=1}^{j} F_k)(\prod_{k=j}^{K} B_k)(1 - M_{ij}) / \prod_{k=1}^{K} F_k\right]\right)}{K} \\
=& \frac{\sum_{j=1}^{K} \left(\left[\check{\mathbf{f}}_j \circ \check{\mathbf{b}}_j \circ (\mathbf{1}_N - \mathbf{m}_j)(\prod_{k=1}^{j} F_k)(\prod_{k=j}^{K} B_k) / \prod_{k=1}^{K} F_k\right] \mathbf{1}_N^T\right)}{K} \\
=& \frac{\sum_{j=1}^{K} \left(\left[\check{\mathbf{f}}_j \circ \check{\mathbf{b}}_j \circ (\mathbf{1}_N - \mathbf{m}_j) \exp\left(\log(\prod_{k=1}^{j} F_k) + \log(\prod_{k=j}^{K} B_k) - \log(\prod_{k=1}^{K} F_k)\right)\right] \mathbf{1}_N^T\right)}{K} \\
=& \frac{\sum_{j=1}^{K} \left(s_j \left[\check{\mathbf{f}}_j \circ \check{\mathbf{b}}_j \circ (\mathbf{1}_N - \mathbf{m}_j)\right] \mathbf{1}_N^T\right)}{K}
\end{aligned} \tag{7}$$

where

$$s_j = \exp\left(\log(\prod_{k=1}^{j} F_k) + \log(\prod_{k=j}^{K} B_k) - \log(\prod_{k=1}^{K} F_k)\right)$$

and each $s_j$, $\check{f}_j$ and $\check{b}_j$ are calculated using the previous estimate of $\mu$.

### 3 Supplementary Note 3

#### 3.1 Accuracy of NNLS estimates from *target-vs-reference* painting

We also evaluated genome-wide ancestry estimates using Non-negative Least Squares (NNLS) to estimate the admixture of each target individual versus a reference panel (Methods). We used the simulation model extended from Simulation 1 (described in Methods): 5 populations admix into the modern population at generation 3000, and after 3 generations, 500 individuals are sampled.

Due to ChromoPainter's inefficiencies when handling larger reference panels, we confined its application to smaller reference sizes. As shown in Supplementary Fig. 3, the NNLS estimation accuracy for ChromoPainter and SparsePainter remains high and comparable, while SparsePainter can process larger reference sizes.

### References


1. Cho, H., Kim, K. M. & Kim, Y. K. Human proline-rich nuclear receptor coregulatory protein 2 mediates an interaction between mRNA surveillance machinery and decapping complex. *Molecular Cell* **33**, 75–86 (2009).
2. Cho, H. *et al.* Glucocorticoid receptor interacts with PNRC2 in a ligand-dependent manner to recruit UPF1 for rapid mRNA degradation. *Proceedings of the National Academy of Sciences* **112**, E1540–E1549 (2015).
3. Zhou, D. & Chen, S. PNRC2 is a 16 kDa coactivator that interacts with nuclear receptors through an SH3-binding motif. *Nucleic Acids Research* **29**, 3939–3948 (2001).
4. Liu, F. *et al.* SRSF10-mediated IL1RAP alternative splicing regulates cervical cancer oncogenesis via mIL1RAP-NF-$\kappa$B-CD47 axis. *Oncogene* **37**, 2394–2409 (2018).
5. URL https://www.proteinatlas.org/ENSG00000148483-TMEM236.
6. Maurya, N. S., Kushwaha, S., Chawade, A. & Mani, A. Transcriptome profiling by combined machine learning and statistical R analysis identifies TMEM236 as a potential novel diagnostic biomarker for colorectal cancer. *Scientific Reports* **11**, 14304 (2021).
7. Adnan Awad, S. *et al.* Mutation accumulation in cancer genes relates to nonoptimal outcome in chronic myeloid leukemia. *Blood Advances* **4**, 546–559 (2020).
8. Yang, L. *et al.* Construction of a predictive model for immunotherapy efficacy in lung squamous cell carcinoma based on the degree of tumor-infiltrating immune cells and molecular typing. *Journal of Translational Medicine* **20**, 1–21 (2022).
9. Rasmussen, A. H., Rasmussen, H. B. & Silahtaroglu, A. The DLGAP family: neuronal expression, function and role in brain disorders. *Molecular Brain* **10**, 1–13 (2017).
10. Taylor, K. M. & Nicholson, R. I. The LZT proteins; the LIV-1 subfamily of zinc transporters. *Biochimica et Biophysica Acta (BBA)-Biomembranes* **1611**, 16–30 (2003).
11. Scarr, E. *et al.* Increased cortical expression of the zinc transporter SLC39A12 suggests a breakdown in zinc cellular homeostasis as part of the pathophysiology of schizophrenia. *npj Schizophrenia* **2**, 1–7 (2016).
12. Davis, D. N. *et al.* A role for zinc transporter gene SLC39A12 in the nervous system and beyond. *Gene* **799**, 145824 (2021).
13. Pipal, K. V. *et al.* Susceptibility loci for type 2 diabetes in the ethnically endogamous Indian Sindhi population: A pooled blood genome-wide association study. *Genes* **13**, 1298 (2022).
14. URL https://www.proteinatlas.org/ENSG00000125900-SIRPD/tissue+cell+type.

15. Tang, J. *et al.* A frameshift variant in the SIRPB1 gene confers susceptibility to Crohn's disease in a Chinese population. *Frontiers in Genetics* **14**, 1130529 (2023).
16. Santiago-Sim, T. *et al.* THSD1 (thrombospondin type 1 domain containing protein 1) mutation in the pathogenesis of intracranial aneurysm and subarachnoid hemorrhage. *Stroke* **47**, 3005–3013 (2016).
17. Conrad, J. A. *et al.* Dominant clonotypes within HIV-specific T cell responses are programmed death-1high and CD127low and display reduced variant cross-reactivity. *The Journal of Immunology* **186**, 6871–6885 (2011).
18. Clark, F. *et al.* Cross-reactivity influences changes in human influenza A virus and Epstein Barr virus specific CD8 memory T cell receptor alpha and beta repertoires between young and old. *Frontiers in Immunology* **13**, 1011935 (2023).
19. Miles, J. J., Douek, D. C. & Price, D. A. Bias in the $\alpha\beta$ T-cell repertoire: implications for disease pathogenesis and vaccination. *Immunology and Cell Biology* **89**, 375–387 (2011).
20. Eliyahu, S. *et al.* Antibody repertoire analysis of hepatitis C virus infections identifies immune signatures associated with spontaneous clearance. *Frontiers in Immunology* **9**, 3004 (2018).
21. Zhuo, Y. *et al.* Evaluation and comparison of adaptive immunity through analyzing the diversities and clonalities of T-cell receptor repertoires in the peripheral blood. *Frontiers in Immunology* **13**, 916430 (2022).
22. Rowntree, L. C. *et al.* A shared TCR bias toward an immunogenic EBV epitope dominates in HLA-B* 07: 02–expressing individuals. *The Journal of Immunology* **205**, 1524–1534 (2020).
23. Aslan, N. *et al.* Severity of acute infectious mononucleosis correlates with cross-reactive influenza CD8 T-cell receptor repertoires. *MBio* **8**, 10–1128 (2017).
24. Robinson, M. W. *et al.* Tracking TCR$\beta$ sequence clonotype expansions during antiviral therapy using high-throughput sequencing of the hypervariable region. *Frontiers in Immunology* **7**, 131 (2016).
25. Gedda, M. R. *et al.* Longitudinal transcriptional analysis of peripheral blood leukocytes in COVID-19 convalescent donors. *Journal of Translational Medicine* **20**, 587 (2022).
26. Lepore, M. *et al.* Parallel T-cell cloning and deep sequencing of human MAIT cells reveal stable oligoclonal TCR$\beta$ repertoire. *Nature Communications* **5**, 3866 (2014).
27. Peng, W. *et al.* Profiling the TRB and IGH repertoire of patients with H5N6 Avian Influenza Virus Infection by high-throughput sequencing. *Scientific Reports* **9**, 7429 (2019).
28. Van de Sandt, C. *et al.* Challenging immunodominance of influenza-specific CD8+ T cell responses restricted by the risk-associated HLA-A* 68: 01 allomorph. *Nature Communications* **10**, 5579 (2019).
29. Fu, Y. *et al.* A comprehensive immune repertoire study for patients with pulmonary tuberculosis. *Molecular Genetics & Genomic Medicine* **7**, e00792 (2019).
30. Mariotti-Ferrandiz, E. *et al.* A TCR$\beta$ repertoire signature can predict experimental cerebral

malaria. *PLoS One* **11**, e0147871 (2016).
31. Yang, J. *et al.* Profiling the repertoire of T-cell receptor beta-chain variable genes in peripheral blood lymphocytes from subjects who have recovered from acute hepatitis B virus infection. *Cellular & Molecular Immunology* **11**, 332–342 (2014).
32. Corpas, M. *et al.* Genetic signature detected in T cell receptors from patients with severe COVID-19. *Iscience* **26** (2023).
33. Liu, J. *et al.* The peripheral differentiation of human natural killer T cells. *Immunology and Cell Biology* **97**, 586–596 (2019).
34. Dong, L., Li, P., Oenema, T., McClurkan, C. L. & Koelle, D. M. Public TCR use by herpes simplex Virus-2–specific human CD8 CTLs. *The Journal of Immunology* **184**, 3063–3071 (2010).
35. Sinha, S. *et al.* An autoimmune disease risk SNP, rs2281808, in SIRPG is associated with reduced expression of SIRP$\gamma$ and heightened effector state in human CD8 T-cells. *Scientific Reports* **8**, 15440 (2018).
36. Zhang, H., Duan, H. O., Kirley, S. D., Zukerberg, L. R. & Wu, C.-L. Aberrant splicing of cables gene, a CDK regulator, in human cancers. *Cancer Biology & Therapy* **4**, 1211–1215 (2005).
37. Sakamoto, H. *et al.* The Cables gene on chromosome 18q is silenced by promoter hypermethylation and allelic loss in human colorectal cancer. *The American Journal of Pathology* **171**, 1509–1519 (2007).
38. Messina, J. P. *et al.* Global spread of dengue virus types: mapping the 70 year history. *Trends in Microbiology* **22**, 138–146 (2014).
39. Seuter, S., Ryynänen, J. & Carlberg, C. The ASAP2 gene is a primary target of 1, 25-dihydroxyvitamin D3 in human monocytes and macrophages. *The Journal of Steroid Biochemistry and Molecular Biology* **144**, 12–18 (2014).
40. Fujii, A. *et al.* The novel driver gene ASAP2 is a potential druggable target in pancreatic cancer. *Cancer Science* **112**, 1655–1668 (2021).
41. Ma, X.-L. *et al.* ASAP2 interrupts c-MET-CIN85 interaction to sustain HGF/c-MET-induced malignant potentials in hepatocellular carcinoma. *Experimental Hematology & Oncology* **12**, 38 (2023).
42. Kim, Y.-g. *et al.* Whole-genome sequencing in clinically diagnosed Charcot–Marie–Tooth disease undiagnosed by whole-exome sequencing. *Brain Communications* **5**, fcad139 (2023).
43. Flynn, E. K. *et al.* Comprehensive Analysis of Pathogenic Deletion Variants in F anconi Anemia Genes. *Human Mutation* **35**, 1342–1353 (2014).
44. Sundaram, A. *et al.* Substrate-driven assembly of a translocon for multipass membrane proteins. *Nature* **611**, 167–172 (2022).
45. McGilvray, P. T. *et al.* An ER translocon for multi-pass membrane protein biogenesis. *Elife* **9**,

e56889 (2020).

46. Guan, Y., Li, Y., Yang, Q.-B., Yu, J. & Qiao, H. LncRNA ABCC6P1 promotes proliferation and migration of papillary thyroid cancer cells via Wnt/$\beta$-catenin signaling pathway. *Annals of Translational Medicine* **9** (2021).
47. Rousseaux, S. *et al.* Ectopic activation of germline and placental genes identifies aggressive metastasis-prone lung cancers. *Science Translational Medicine* **5**, 186ra66–186ra66 (2013).
48. Lawlor, M. A. *et al.* Essential role of PDK1 in regulating cell size and development in mice. *The EMBO Journal* **21**, 3728–3738 (2002).
49. Nalairndran, G. *et al.* Phosphoinositide-dependent Kinase-1 (PDPK1) regulates serum/glucocorticoid-regulated Kinase 3 (SGK3) for prostate cancer cell survival. *Journal of Cellular and Molecular Medicine* **24**, 12188–12198 (2020).
50. Luo, D. *et al.* The PDK1/c-Jun pathway activated by TGF-$\beta$ induces EMT and promotes proliferation and invasion in human glioblastoma. *International Journal of Oncology* **53**, 2067–2080 (2018).
51. Maurer, M. *et al.* 3-phosphoinositide–dependent kinase 1 potentiates upstream lesions on the phosphatidylinositol 3-kinase pathway in breast carcinoma. *Cancer Research* **69**, 6299–6306 (2009).
52. Ahmed, N., Riley, C. & Quinn, M. An immunohistochemical perspective of PPAR$\beta$ and one of its putative targets PDK1 in normal ovaries, benign and malignant ovarian tumours. *British Journal of Cancer* **98**, 1415–1424 (2008).
53. Pietri, M. *et al.* PDK1 decreases TACE-mediated $\alpha$-secretase activity and promotes disease progression in prion and Alzheimer's diseases. *Nature Medicine* **19**, 1124–1131 (2013).
54. Sun, Z. *et al.* The kinase PDK1 is critical for promoting T follicular helper cell differentiation. *Elife* **10**, e61406 (2021).
55. Ueta, M. *et al.* Genome-wide association study using the ethnicity-specific Japonica array: identification of new susceptibility loci for cold medicine-related Stevens–Johnson syndrome with severe ocular complications. *Journal of Human Genetics* **62**, 485–489 (2017).
56. Susceptibility genes and HLA for cold medicine-related SJS/TEN with SOC, author=Ueta, Mayumi. *Frontiers in Genetics* **13**, 912478 (2022).
57. Tian, C. *et al.* Genome-wide association and HLA region fine-mapping studies identify susceptibility loci for multiple common infections. *Nature Communications* **8**, 599 (2017).
58. Herold, C. *et al.* Family-based association analyses of imputed genotypes reveal genome-wide significant association of Alzheimer's disease with OSBPL6, PTPRG, and PDCL3. *Molecular Psychiatry* **21**, 1608–1612 (2016).
59. Tournel, G. *et al.* Molecular analysis of the CYP2F1 gene: identification of a frequent non-functional allelic variant. *Mutation Research/Fundamental and Molecular Mechanisms of Mutagenesis* **617**, 79–89 (2007).

60. Korytina, G., Akhmadishina, L. & Viktorova, T. The CYP1B1 and CYP2F1 genes polymorphisms frequency in three ethnic groups of Bashkortostan and chronic obstructive pulmonary disease patients. *Molekuliarnaia Biologiia* **44**, 33–41 (2010).
61. Androutsopoulos, V. P., Tsatsakis, A. M. & Spandidos, D. A. Cytochrome P450 CYP1A1: wider roles in cancer progression and prevention. *BMC Cancer* **9**, 1–17 (2009).
62. Jiang, J. H. *et al.* Study on genetic polymorphisms of CYP2F1 gene in Guangdong population of China. *Chinese Journal of Medical Genetics* **23**, 383–387 (2006).
63. Seo, S. *et al.* BBS6, BBS10, and BBS12 form a complex with CCT/TRiC family chaperonins and mediate BBSome assembly. *Proceedings of the National Academy of Sciences* **107**, 1488–1493 (2010).
64. Hirayama, S. *et al.* MKKS is a centrosome-shuttling protein degraded by disease-causing mutations via CHIP-mediated ubiquitination. *Molecular Biology of the Cell* **19**, 899–911 (2008).
65. Slavotinek, A. M. *McKusick-Kaufman Syndrome* (University of Washington, Seattle, Seattle (WA), 1993).
66. Robinson, N. J. *et al.* SLX4IP and telomere dynamics dictate breast cancer metastasis and therapeutic responsiveness. *Life Science Alliance* **3** (2020).
67. Meissner, B. *et al.* Frequent and sex-biased deletion of SLX4IP by illegitimate V (D) J-mediated recombination in childhood acute lymphoblastic leukemia. *Human Molecular Genetics* **23**, 590–601 (2014).
68. Urnikyte, A., Masiulyte, A., Pranckeniene, L. & Kučinskas, V. Disentangling archaic introgression and genomic signatures of selection at human immunity genes. *Infection, Genetics and Evolution* **116**, 105528 (2023).
69. Watterson, G. On the number of segregating sites in genetical models without recombination. *Theoretical Population Biology* **7**, 256–276 (1975).

### Supplementary Figures

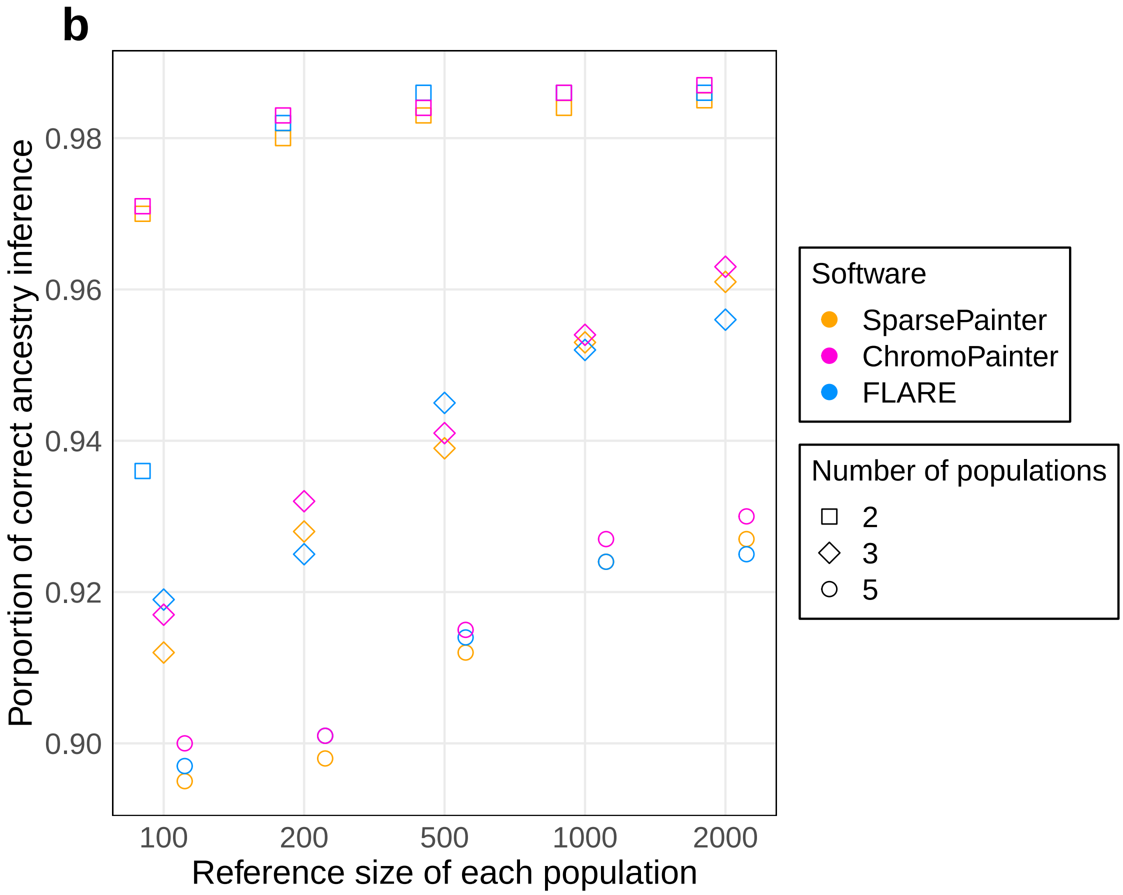


**Supplementary Fig. 1: Proportion of correct ancestry inference of local ancestry estimates when painting target individuals with different reference panels.** This simulation has 20000 SNPs and 50 target individuals sampled 13 generations after admixture. The y-axis is the proportion for correct local ancestry inference, and the x-axis is the reference size of each reference ancestry.

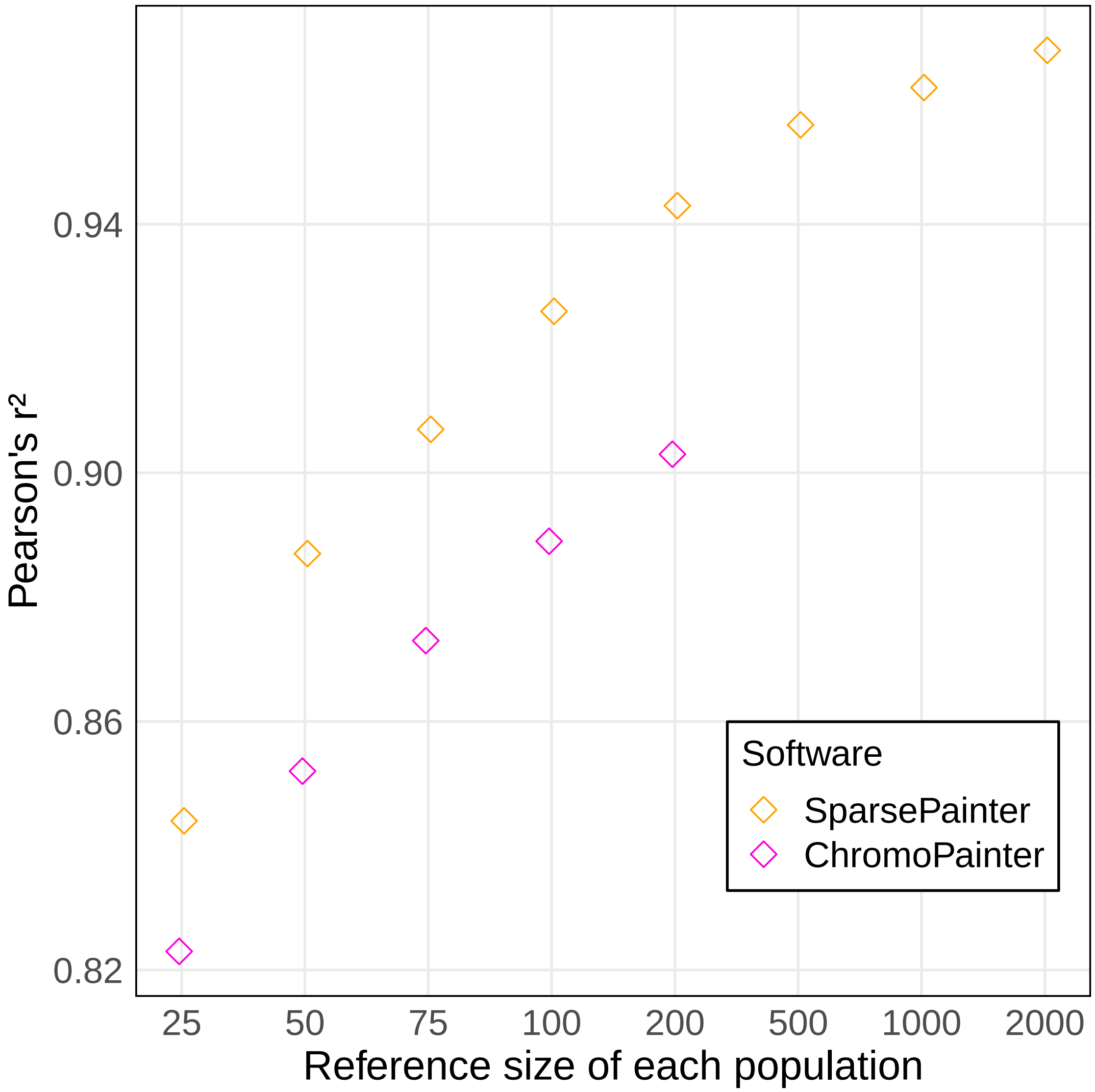


**Supplementary Fig. 2**: **Accuracy of NNLS estimates for *target-vs-reference* painting.** This simulation has 20000 SNPs and shows that SparsePainter and ChromoPainter have comparable accuracy.

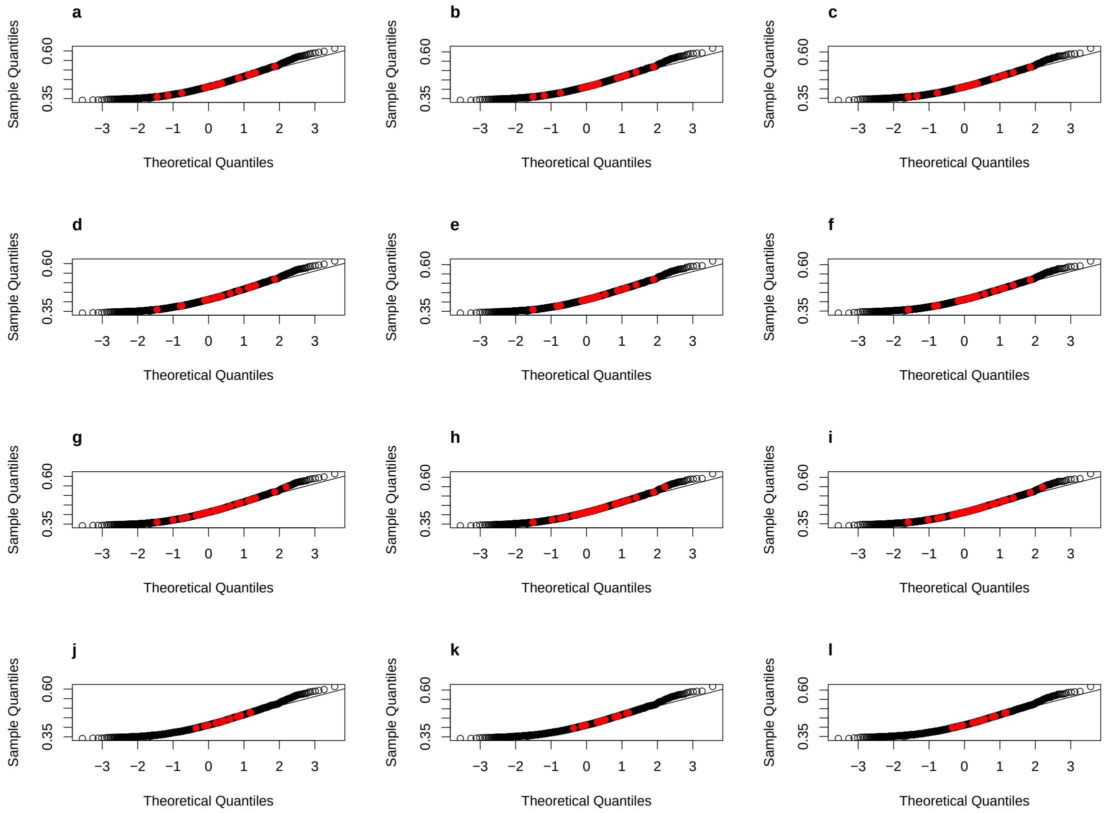


**Supplementary Fig. 3**: **QQ plots for the frequency of G+C in Africa, Europe and East Asia.** The red points represent the regions with shared LDAS signals in 26-pop painting (a,b,c) and 5-continent painting (d,e,f), and shared AAS signals in 26-pop painting (g,h,i) and 5-continent painting (j,k,l). a,d,g,j are the QQ-plots for the frequency of G+C in Africa; b,e,h,k are the QQ-plots for the frequency of G+C in Europe; c,f,i,l are the QQ-plots for the frequency of G+C in East Asia.

### Supplementary Tables

| Selection Type | Genes |
|---|---|
| Ethnicity-specific AAS signals | **both 26-pop and 5-continent painting:** 1:RNU5E-1, 1:KIAA2013, 1:PLOD1, 1:LINC01781, 1:GSTM1, 1:GSTM2, 2:SCHLAP1, 3:LRIG1, 3:ROBO2, 6:H2BC6, 6:H1-12P, 6:H2AC7, 6:H4C5, 6:LINC03004, 6:LINC02539, 8:DLGAP2, 9:ADGRD2, 9:NR5A1, 21:WDR4 |
| | **26-pop painting only:** 1:C1orf167, 1:MTHFR, 1:CLCN6, 1:NPPA, 1:MFN2, 1:MYOM3, 1:MYOM3-AS1, 1:LINC01758, 1:AMPD2, 1:GSTM4, 1:GSTM5, 1:GSTM3, 1:EPS8L3, 1:FCGR3A, 1:FCGR2C, 1:FCGR3B, 1:OR2G6, 1:OR2T11, 2:ITGB1BP1, 2:CPSF3, 2:IAH1, 2:ADAM17, 2:AGAP1, 3:CACNA2D3, 4:KCNIP4, 5:DNAH5, 6:H2BC8, 6:H3C5P, 6:H2AC9P, 6:H1-3, 6:H4C6, 6:H4C7, 6:HBS1L, 6:MYB, 6:IL20RA, 7:GSDME, 7:OSBPL3, 7:IRF5, 7:LUC7L2, 7:FMC1-LUC7L2, 8:CSMD1, 8:SGCZ, 8:ADCY8, 8:PARP10, 8:SPATC1, 8:OPLAH, 9:MFSD14B, 9:NR6A1, 10:ADARB2, 10:ADARB2-AS1, 15:PWRN4, 15:PWRN1, 15:PPIP5K1, 15:CKMT1B, 15:STRC, 15:NEDD4, 15:CNOT6LP1, 15:LINC00927, 16:ERVK13-1, 16:RPS15A, 16:ARL6IP1, 16:SMG1, 16:SMG1-DT, 17:SLFN11, 19:MUC16, 19:ZNF568, 20:BFSP1, 21:ANKRD20A11P, 21:RUNX1 |

**Supplementary Tab. 1**: **Genes of ethnicity-specific AAS signals across 7 UK Biobank self-reported ethnic backgrounds.** Display Format: Chromosome:Gene.

**Supplementary Tab. 2: Summary statistics for 10,000 UK Biobank individuals with self-reported British ethnic background at each SNP, including LDAS, AAS, and average probabilities of 26 1000GP populations.** The LDAS of SNPs which are removed in quality control are displayed as ‘NA’.

**Supplementary Tab. 3: Summary statistics for 12,713 UK Biobank individuals with self-reported Irish ethnic background at each SNP, including LDAS, AAS, and average probabilities of 26 1000GP populations.** The LDAS of SNPs which are removed in quality control are displayed as ‘NA’.

**Supplementary Tab. 4: Summary statistics for 3,203 UK Biobank individuals with self-reported African ethnic background at each SNP, including LDAS, AAS, and average proba-**

**bilities of 26 1000GP populations.** The LDAS of SNPs which are removed in quality control are displayed as 'NA'.

**Supplementary Tab. 5: Summary statistics for 4,279 UK Biobank individuals with self-reported Caribbean ethnic background at each SNP, including LDAS, AAS, and average probabilities of 26 1000GP populations.** The LDAS of SNPs which are removed in quality control are displayed as 'NA'.

**Supplementary Tab. 6: Summary statistics for 5,660 UK Biobank individuals with self-reported Indian ethnic background at each SNP, including LDAS, AAS, and average probabilities of 26 1000GP populations.** The LDAS of SNPs which are removed in quality control are displayed as 'NA'.

**Supplementary Tab. 7: Summary statistics for 1,747 UK Biobank individuals with self-reported Pakistani ethnic background at each SNP, including LDAS, AAS, and average probabilities of 26 1000GP populations.** The LDAS of SNPs which are removed in quality control are displayed as 'NA'.

**Supplementary Tab. 8: Summary statistics for 1,503 UK Biobank individuals with self-reported Chinese ethnic background at each SNP, including LDAS, AAS, and average probabilities of 26 1000GP populations.** The LDAS of SNPs which are removed in quality control are displayed as 'NA'.

**Supplementary Tab. 9: SNPs with shared LDAS or AAS signals.** The structural variation information in 1000GP for those SNPs reported by Sudmant et al. (2015) is also listed.
